# Humoral and cellular immune responses against SARS-CoV-2 variants and human coronaviruses after single BNT162b2 vaccination

**DOI:** 10.1101/2021.04.16.21255412

**Authors:** Metodi V. Stankov, Anne Cossmann, Agnes Bonifacius, Alexandra Dopfer-Jablonka, Gema Morillas Ramos, Nina Gödecke, Anna Zychlinsky Scharff, Christine Happle, Anna-Lena Boeck, Anh Thu Tran, Isabell Pink, Marius M. Hoeper, Rainer Blasczyk, Martin S. Winkler, Inga Nehlmeier, Amy Kempf, Heike Hofmann-Winkler, Markus Hoffmann, Britta Eiz-Vesper, Stefan Pöhlmann, Georg M.N. Behrens

**Author notes:** **Corresponding author:** Professor Georg M.N. Behrens, Department of Rheumatology and Immunology, Hannover Medical School, Carl-Neuberg-Straße 1, D - 30625 Hannover, Germany.

## Abstract

Vaccine-induced neutralizing antibodies are key in combating the COVID-19 pandemic. However, delays of boost immunization due to limited availability of vaccines may leave individuals vulnerable to infection and disease for prolonged periods. The emergence of SARS-CoV-2 variants of concern (VOC), B.1.1.7 (United Kingdom), B.1.351 (South Africa) and P.1 (Brazil), may reinforce this issue with the latter two being able to evade control by antibodies. We assessed humoral and T cell responses against SARS-CoV-2 WT and VOC and endemic human coronaviruses (hCoV) that were induced after single and double vaccination with BNT162b2. Despite readily detectable IgG against the receptor-binding domain (RBD) of the SARS-CoV-2 S protein at day 14 after a single vaccination, inhibition of SARS-CoV-2 S-driven host cell entry was weak and particularly low for the B.1.351 variant. Frequencies of SARS-CoV-2 specific T cells were low in many vaccinees after application of a single dose and influenced by immunity against endemic hCoV. The second vaccination significantly boosted T cell frequencies reactive for WT, B.1.1.7 and B.1.351 variants. These results call into question whether neutralizing antibodies significantly contribute to protection against COVID-19 upon single vaccination and suggest that cellular immunity is central for the early defenses against COVID-19.

## Introduction

Several vaccines encoding the viral spike (S) protein have been approved to combat the coronavirus disease 2019 (COVID-19) pandemic (1-3). The recent emergence of the severe acute respiratory syndrome coronavirus 2 (SARS-CoV-2) variants of concern (VOC) B.1.1.7 in the UK, B.1.351 in South Africa, and P.1 in Brazil might threaten measures to control the COVID-19 pandemic due to their ease of transmission (4, 5) and, in case of variants B.1.351 and P.1, resistance to neutralization by monoclonal antibodies (mAbs) and partial resistance to neutralization by antibodies induced upon infection and vaccination (6-10).

Results from clinical trials suggest that vaccine protection against COVID-19 begins around two weeks after the first vaccine dose (1, 2). However, only modest neutralization activity of sera was observed shortly before the second vaccine administration, and robust increase in neutralizing antibody titers required a second boosting dose (11, 12). Due to the accelerating pandemic and the associated need to provide at least partial protection at the population level, the U.K. Joint Committee on Vaccines and Immunization has proposed extending the time to the second vaccine dose to enable first vaccination of more individuals within a short time period (13).

However, delaying time until the second vaccination may lead to a sizable population of vaccinees with incomplete or short-lived anti-SARS-CoV-2 immunity and this approach may even favor the emergence of escape variants. In order to address this question, we analyzed cellular and humoral immune responses induced by a single dose vaccination of the mRNA vaccine BNT162b2. We also determined the impact of preexisting immunity against human coronavirus (hCoV) on the vaccine response.

## Results and discussion

Anti-SARS-CoV-2 S IgG and IgA levels were determined in individuals early (mean 8.7 days, range 2 to 14 days) and late (mean 20.6 days, range 17-27 days) after immunization with a single 30 µg dose of BNT162b2 (n=124). In addition, samples obtained at mean 21 days (range 6-36 days) after a second 30 µg dose (n=69) were analyzed. Antibodies of the IgG subtype directed against the S1 subunit of SARS-CoV-2 S became detectable around day 14 after the first shot, with almost all participants having measurable IgG levels 17 days after the first BNT162b2 dose (Fig. 1 A-B). Anti-SARS-CoV-2 IgA (n= 54) was detectable in all individuals at a mean of 20.2 days (range 19-25 days) after the first vaccination (Fig. 1C). The magnitude of the anti-SARS-CoV-2 S IgG antibody response was significantly higher 21 days after the second BNT162b2 dose (Fig. 1B).

**Figure 1:**
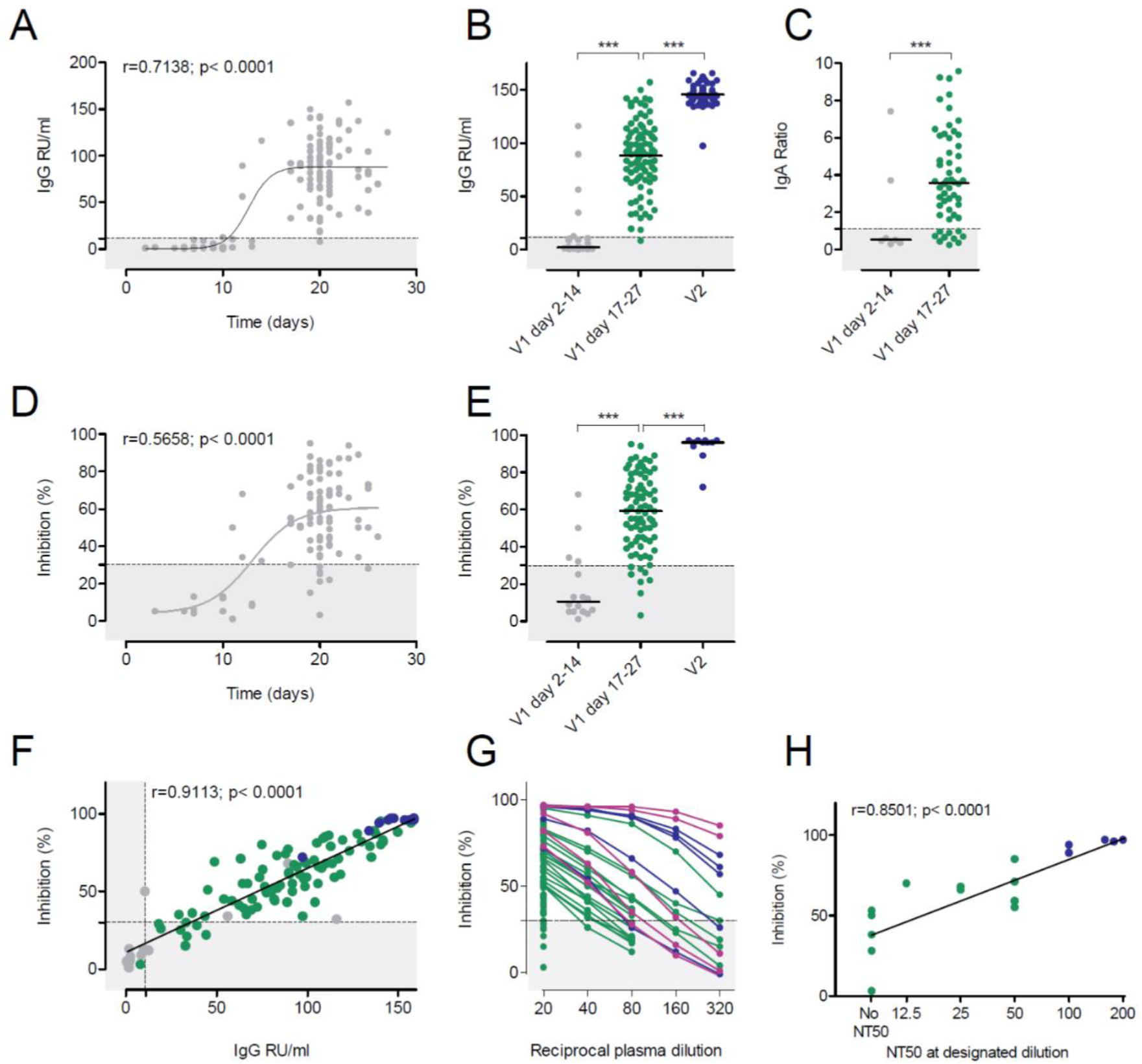
Humoral immune response after BNT162b2 vaccination. **(A)** Time course of anti-SARS-CoV-2 S protein IgG (n=121) after the first BNT162b2 dose. **(B)** Anti-SARS-CoV-2 S IgG responses in relative units per mL (RU/mL) after the first (V1, grey and green dots, n=32 and n=92, respectively) and second (V2, blue dots, n=69) BNT162b2 dose as indicated. **(C)** Anti-SARS-CoV-2 S IgA responses (n=63) after the first BNT162b2 vaccination. ELISA results are depicted as sample/calibrator ratio. **(D)** Time course of percent inhibition in the sVNT after the first BNT162b2 dose (n=99). **(E)** Inhibition in the sVNT after the first or second BNT162b2 vaccination as indicated (grey dots, n=16, green dots, n=84, blue dots, n=11). **(F)** Correlation of inhibition (sVNT) and anti-SARS-CoV-2 S IgG (ELISA) after the first or second BNT162b2 vaccination as indicated for E. **(G)** Reciprocal plasma dilutions of the sVNT in convalescent COVID-19 patients (purple), after the second BNT162b2 dose (blue) or after the first BNT162b2 dose (green). Note that only samples >50% inhibition at the lowest dilution (1:20) were further titrated. **(H)** Correlation of the sVNT inhibition with the NT50 of the pseudotype virus neutralization results (n=17) as indicated. Dotted lines represent the assay cut offs as suggested by the manufacturer. ***p<0.001 by two-tailed student’s t test or ANOVA with Bonferroni’s post hoc analysis, where appropriate.

When testing plasma samples in a surrogate virus neutralization test (sVNT) for inhibition of RBD binding to plate-bound ACE2 receptor, a similar picture emerged (Fig. 1D-E). Most plasma samples from day 2 to 14 after the first BNT162b2 vaccination remained below the cut-off (30%) of the assay. In contrast, almost all participants had anti-SARS-CoV-2 S1 RBD inhibitory antibodies detectable beyond day 17 after first BNT162b2 vaccination. The second vaccination significantly increased inhibitory activity in this assay. The sVNT showed a highly statistically significant correlation to inhibition of SARS-CoV-2 S-driven host cell entry in a vesicular stomatitis virus (VSV)-pseudotype-based assay for detection of neutralizing antibody responses (Fig. 1F). To further assess the inhibitory activity of plasma samples 17-21 days after the first BNT162b2, we diluted plasma with >50 % inhibition in the sVNT and compared the results to those from convalescent COVID-19 patients or individuals 21 days after the second BNT162b2 vaccination. Plasma samples with inhibitory activity less than 90 % at the highest plasma concentration (1:20) showed a rapid and linear decline by dilution. Only samples with baseline inhibition > 90% maintained > 50% inhibition in the sVNT upon further dilution (Fig. 1G) indicating low antibody concentrations in most plasma samples. Our data support the finding that SARS-CoV-2 need little affinity maturation (14, 15), and become detectable in the plasma at 10-14 days post first vaccination.

We next determined whether antibodies induced by a single BNT162b2 vaccination inhibited host cell entry driven by WT S protein (harboring D614G) and the S proteins of variants B.1.1.7, B.1.351 and P.1. For this, we used a VSV-based vector pseudotyped with respective S proteins, as previously described (8). Plasma collected from patients with severe and current COVID-19 due to SARS-CoV-2 WT was included as control. These plasma samples reduced entry driven by WT S and the S protein of variant B.1.1.7 with similar efficiency (Fig. 2A). In contrast, blockade of entry driven by the S protein of P.1 and particularly the B.1.351 variant was less efficient (Fig. 2A), which is consistent with our published data (8). Similarly, and again in line with our previous results (8), plasma collected from vaccinees 21 days after the second BNT162b2 dose efficiently neutralized entry driven by the WT S protein and inhibition of entry driven by the S protein of B.1.1.7 was only marginally reduced (Fig. 2C). In contrast, inhibition of entry driven by the S proteins of variants P.1 and particularly B.1.351 was less efficient (Fig. 2C). Finally, plasma samples from the same donors obtained 21 days after the first dose exerted no (n = 3) or low (n = 2) inhibitory activity and reduced inhibition of entry driven by the S protein of B.1.351 was observed (Fig. 2B). The overall summary of the inhibition analysis of n=14 vaccinees after a single dose is depicted in Fig. 2D and suggests that a single vaccination might frequently fail to induce a measurable neutralizing antibody response. Moreover, if such a response is induced, it may fail to protect against infection with the B.1.351 variant.

**Figure 2:**
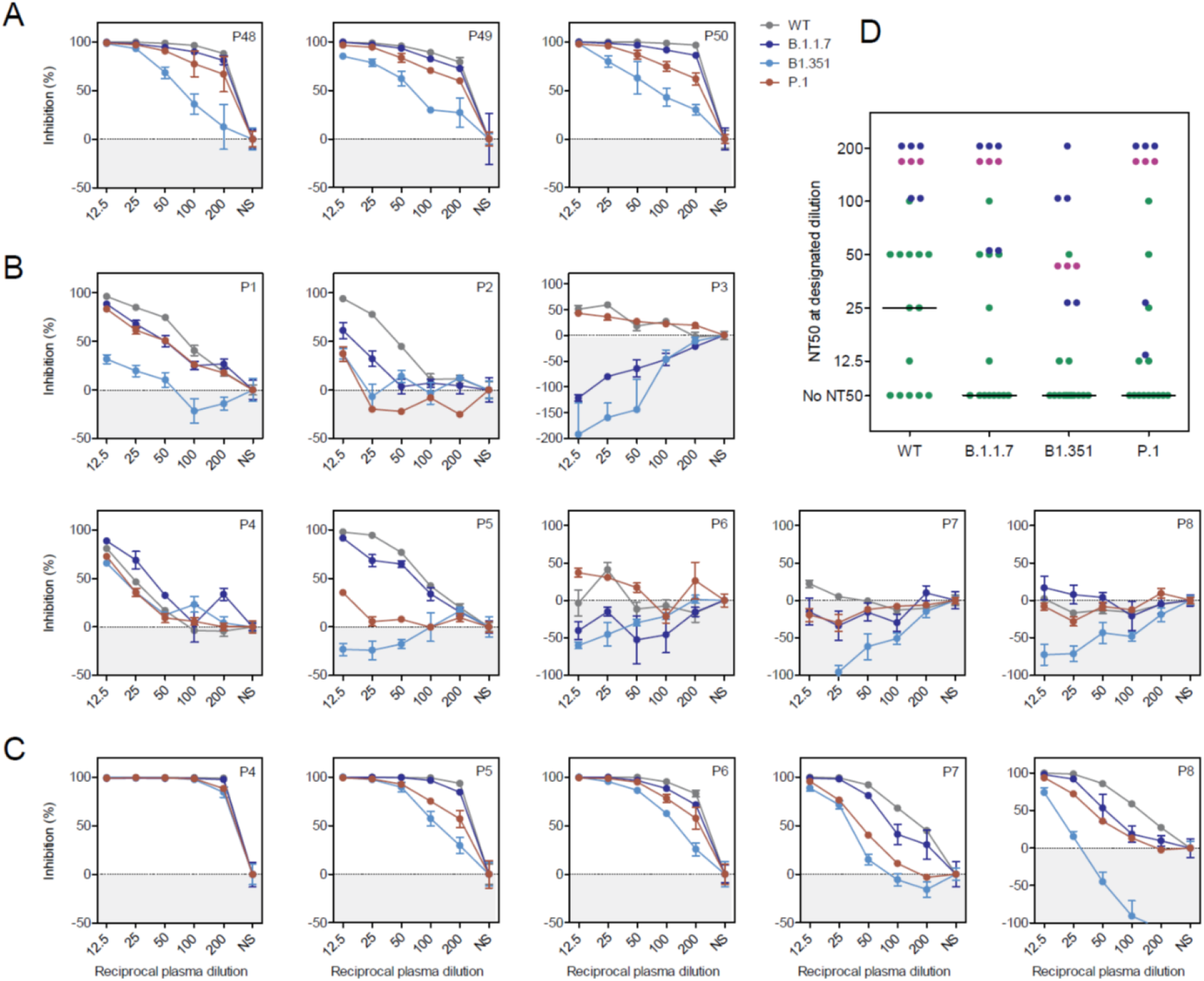
S protein of SARS-CoV-2 WT, B1.1.7, B.1351, and P.1 variants show reduced neutralization sensitivity against plasma from single and twice BNT162b2 vaccinated individuals. Pseudotypes bearing the indicated S proteins were incubated with different dilutions of plasma derived from patients with severe COVID-19 **(A)** or plasma obtained shortly before the second dose BNT162b2 **(B)** or 21 days after the second dose **(C)** and inoculated onto Vero target cells. Transduction efficiency was quantified by measuring virus-encoded luciferase activity in cell lysates at 16-20 h post transduction. The results are shown as % inhibition. For normalization, S protein-driven entry in the absence of plasma was set as 0%. Presented are the data from a single experiment performed with technical triplicates. Error bars indicate SD. Most results were confirmed in a second biological replicate. **(D)** Plasma dilutions that lead to a 50 % reduction in S protein-driven transduction (neutralization titer 50, NT50) were calculated for convalescent COVID-19 plasma (purple, n=3) and vaccinee plasma after the first (green, n=14) and second BNT162b2 dose (blue, n=5). The line represents the median NT50 of single vaccinated individuals. WT = wildtype, NS = no serum.

Besides neutralizing antibodies, the S protein also harbors T-cell epitopes which are central in COVID-19 immunity (16, 17). To assess T cell immunity post vaccination, we analyzed the frequencies of T cells producing interferon-gamma (IFNγ) upon stimulation with peptide pools derived from the S protein of SARS-CoV-2, hCoV-OC43 and hCoV-299E, and cytomegalovirus (CMV) pp65 (as control) by enzyme-linked immunospot assay (EliSpot). T cells reactive to peptide stimulation from SARS-CoV-2 WT, B.1.1.7, and B.1.351 were undetectable in more than 40% of vaccinees after a single BNT162b2 shot (Fig. 3A-C) but increased significantly following boosting (Fig. 3C). Using an alternative *in vitro* SARS-CoV-2 specific cytokine release assay analogous to the tuberculosis IFNγ release assay (18, 19), we observed significantly increased IFNγ production by PBMCs after the first and second BNT162b2 vaccination as compared to controls, but responses remained low in a sizable proportion of individuals after only one vaccination (Fig. 3D). Taken together, our data are in line with our previous analyses in convalescent COVID-19 patients (15) and shows that the magnitude of B and T cell responses against SARS-CoV-2 upon vaccination is wide-ranging and differs for distinct virus variants. Particularly, the magnitude of SARS-CoV-2-specific T cell responses shows great heterogeneity and is not readily detectable after a single shot.

**Figure 3:**
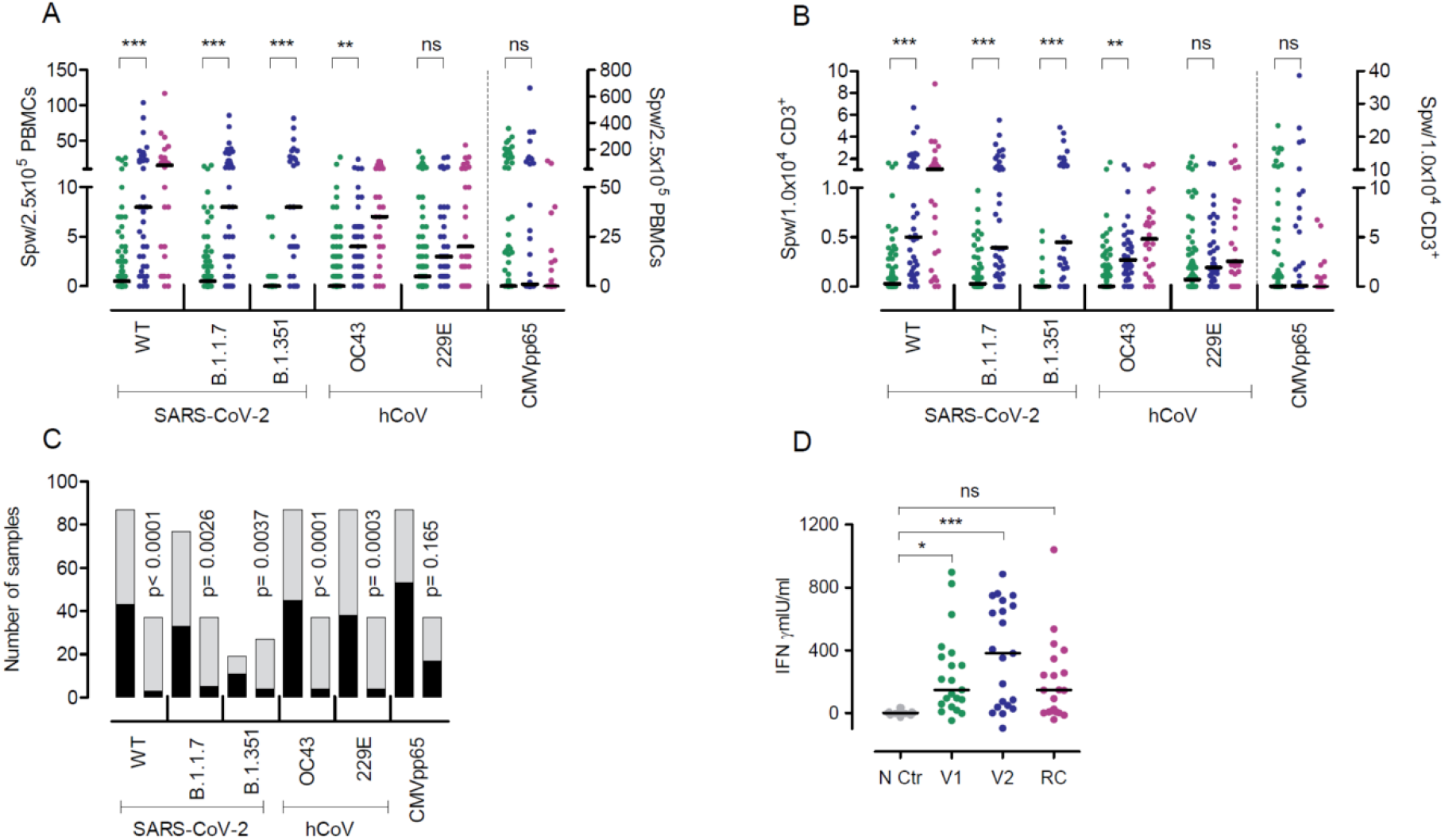
T cell frequencies against SARS-CoV-2 WT and variants B.1.1.7 and B.1.351, and hCoV OC43 and 229E after the first and second BNT162b2 dose. IFNγ EliSpot data for the SARS-CoV-2 variants and two hCoV from individuals vaccinated once (n=78-88) or twice (n=27-37) are shown. The first samples (green dots) depict data from shortly before the second vaccination and the second sample (blue dots) from 21 days after the second BNT162b2 dose. Purple dots represent results from convalescent patients mean 8.5 weeks (range 3-36 weeks) after mild COVID-19. **(A)** Data are depicted as the number of spots per well (spw)/2.5×10^5^ PBMCs or **(B)** as spw/1×10^4^ CD3^+^ T cells. For CMVpp65, values from individuals irrespective if their CMV serostatus are depicted (note the separate axis on the right). **(C)** Number of individuals with zero (black) or ≥1 (grey) spw in the IFNγ EliSpot after the first (left column) or second (right column) BNT162b2 dose. **(D)** IFNγ release assay results obtained from PBMCs of single (green dots) or twice (blue dots) vaccinated individuals after restimulation with SARS-CoV-2 S protein for 24 h and assessment of IFNγ in the supernatant by ELISA. Negative controls (NC, grey dots) are from the same individuals as after the second vaccination (blue dots) but from PBMC collected in 2019 before vaccination. Purple dots represent results from PBMCs of convalesecent patients with mild to severe COVID-19 (RC) at a mean 10.1 weeks (range 7-14 weeks) after symptom onset. Bars represent median. *p<0.05, **p<0.005, *** p<0.001, by two-tailed student’s t test or ANOVA with Bonferroni’s post hoc analysis, where appropriate. P-values in C are calculated by Fisher’s exact test, ns=not significant.

SARS-CoV-2 is a member of the coronavirus family that includes hCoV-OC43, hCoV-HKU1, hCoV-229E, and hCoV-NL63. For the two hCoV variants testes in our work, we observed a significant expansion of hCoV-OC43 reactive T cells and an increase in hCoV-229E responsive T cells in the EliSpot (Fig.3 A+B), suggesting a strong overlap of hCoV with SARS-CoV-2 immunity upon vaccination. This overlap in response was further demonstrated by the significant positive correlation of SARS-CoV-2 WT and variants B.1.1.7 and B.1.351 responsive T cell frequencies with those against hCoVs OC43 and 229E (Fig. 4A+B) after the first and second BNT162b2 vaccination. Also, T cell frequencies for the B.1.1.7 variant correlated closely with hCoV tested in our study (OC43, r=0.6795, p<0.0001, 229E, r=0.646, p<0.0001, *data not shown*). These correlations were observed upon first and second BNT162b2 vaccination, while no correlation between SARS-CoV-2 T cell responses and those towards the unrelated virus CMV occurred (Fig. 4A).

**Figure 4:**
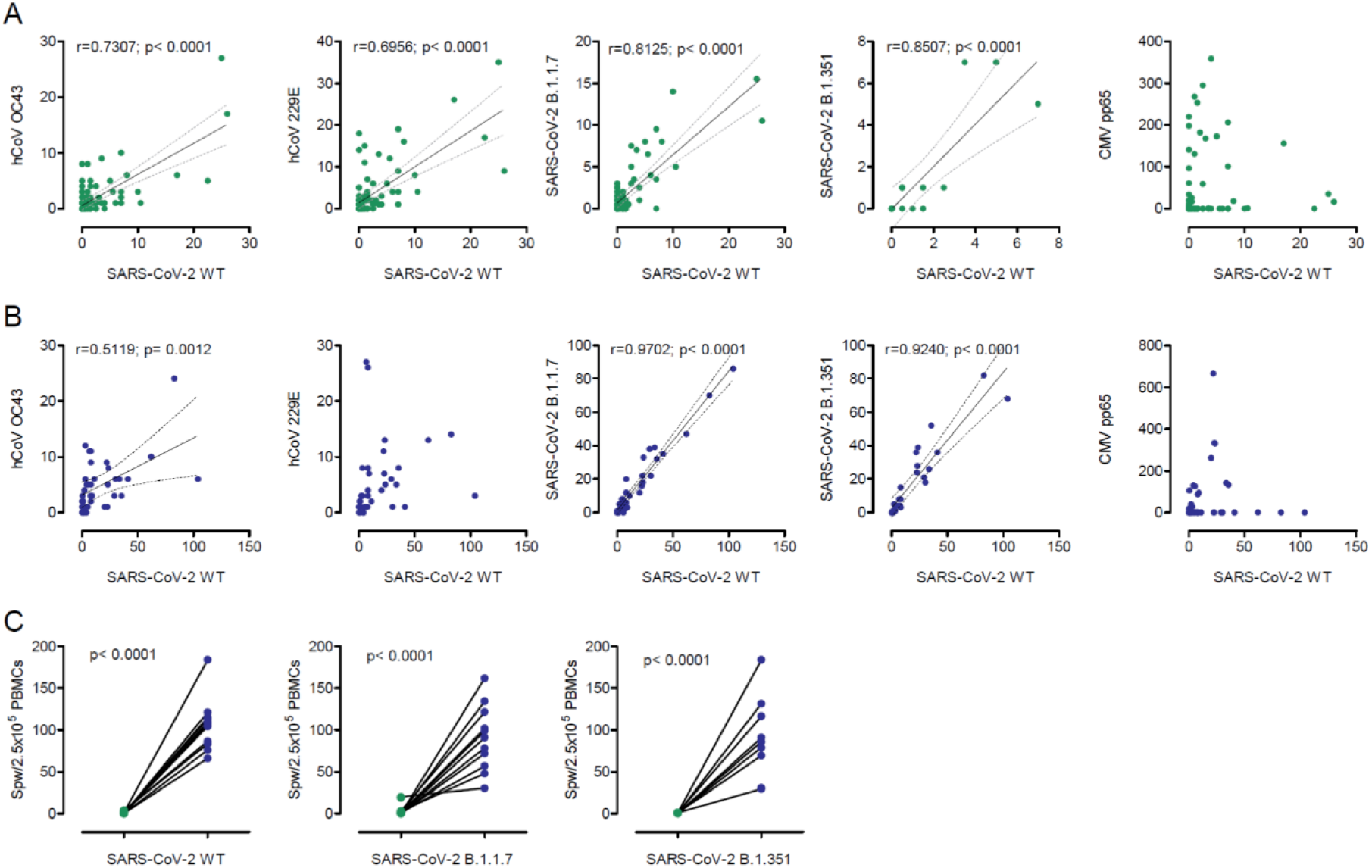
Correlation of T cell immune response against SARS-CoV-2 WT and variants B.1.1.7 or B.1.351, and hCoV OC43 and 229E after a single and double BNT162b2 dose. **(A)** Correlation of IFNγ EliSpot data (depicted as spw/2.5×10^5^ PBMCs) after the first BNT162b2 dose between S protein of SARS-CoV-2 WT, hCoV, VOC, as indicated, and CMVpp65 as controls. **(B)** Same as in A but after the second vaccine dose. Only significant correlations are depicted. **(C)** PBMCs from individuals with low or zero spw after a single BNT162b2 dose (green dots) were *in vitro* stimulated with overlapping peptide pools from SARS-CoV-2 S protein for seven days and again assessed in the IFNγ EliSpot with S protein-derived peptide pools (blue dots) from SARS-CoV-2 WT or variants B.1.1.7 and B.1.351 as indicated. Groups were compared by two-tailed paired student’s t test.

These data on strongly related intraindividual hCoV and SARS-CoV-2 immune responses are in line with our analyses in convalescent COVID-19 patients (20) and previously described associations described overlapping B cell responses against α and β-hCoVs (21). Cross-reactivity against SARS-CoV-2 and endemic hCoVs are mediated primarily by memory CD4+ T cell responses directed against conserved epitopes and have been reported in up to 50% of individuals (15, 16, 22-24). Expectedly, also T cell frequencies against SARS-CoV-2 WT correlated closely and increasingly after the second vaccination with those observed for SARS-CoV-2 VOC (Fig. 4 A+B). Here, we provide initial evidence that cross-reactivity also occurs through COVID-19 vaccination and suggest that individuals with cross-reactive T cells may respond differently to vaccines than those without such memory (25, 26).

Prompted by weak antibody neutralization activity in almost all individuals and low or even undetectable T cell frequencies after the first vaccination, we performed experiments to expand vaccine-induced T cells. For this, we stimulated PBMCs with SARS-CoV-2 S1 and S2 peptide pools from WT and VOC for seven days, which led to expansion and detection of responding T cells even in those individuals with initially low or no T cell response (Fig. 4 C). This suggests efficient T memory cell generation or booster of natural immunity against coronavirus variants after single vaccination, which was reactive after long-term *in vitro* stimulation with WT and mutant SARS-CoV-2 S peptide variants. These results provide evidence for potentially effective, albeit weak, T cell immune responses against SARS-CoV-2 WT and VOC in a relevant proportion of individuals vaccinated with only the initial dose. Studies in convalescent COVID-19 patients have described that efficient SARS-CoV-2-specific T cell responses are associated with milder disease (15, 27), suggesting that T cell responses may be central to control of SARS-CoV-2 infection. However, our study does not allow us to estimate whether these exclusively S-protein specific T cell responses significantly add to protection against COVID-19. Specific correlates of protection can only be established by studies observing a significant number of re-infections over time (15).

Our study is limited by the fact that we were unable to assess T cell responses before vaccination. Secondly, the analyzed cellular responses would benefit from further identification of T cell subsets and viral epitopes involved. Third, our study only considers systemic responses and studies of airway compartments or tissue-resident T cells may be important to gain additional insights into protective immunity after vaccination against COVID-19.

In summary, our data demonstrate suboptimal neutralizing antibody activity against SARS-CoV-2 WT and VOC after a single BNT162b2 vaccination, in keeping with a study deposited on a preprint server (26). T cells, which responded equally to spike-derived peptides from SARS-CoV-2 WT, B.1.1.7 and B.1.351 were detectable with a broad inter-individual range and influenced by cross-reactive T cells against hCoV. We propose that non-neutralizing antibody function and/or cellular immunity constitute an important outcome after vaccination and may be part of the early defense against SARS-CoV-2 infection. We conclude that without an immune correlate of protection for SARS-CoV-2 vaccines, protective immunity after vaccination cannot be precisely measured and variations in effective immunization programs cannot be confidently recommended (26, 28).

## Methods

### Study approval

The study was approved by the Internal Review Board of Hannover Medical School (MHH, approval number 3639_2017, 8973_BO-K_2020, 9226_BO_K_2020, 9255_BO_K_2020, 9459_BO_K_2020) and University Medicine Göttingen (approval number SeptImmun Study 25/4/19 Ü). For other methods, see supplementary material.

## Data Availability

Data are available upon request to the corresponding author.

## Author’s contributions

GMNB and SP designed the research study and experiments, supervised data collection, performed data analysis and interpretation. AD-J and CH designed the study and contributed to data analysis and interpretation. AC and HH-W coordinated sample and data collection, organized biobanking, data management or experiments. AZS, IP, MMH, MSW, A-LB, NG and ATT contributed to sample collection and study organization. MVS and GMR conducted experiments and analyzed data. SP designed research experiments, supervised data collection, performed data analysis and interpretation. MH supervised data collection, performed data analysis and interpretation. IN and AK conducted experiments and acquired data. AB, RB and BE-V designed or performed T cell research experiments, performed data analysis and interpretation. All authors contributed to writing the manuscript.

## Acknowledgments

We thank all participants of the CoCo study. We would like to thank Marion Hitzigrath, Sabine Buyny, Melanie Ignacio, Annika Heidemann, Luis Manthey, Till Redeker, Elisa Armbrecht, Annkathrin Anton, Christian Sturm, Julia Wahlen, Anna Zeisler, Oliver Keil, Mathäus Vetter, Andreas Bode, Birgit Heinisch, Gudrun Mielke, Nele Stein Daniel Gussarow, Juliane Ebersold, Nicole Neumann, Dörthe Rokitta, and Sophie Meyer for technical and logistical support. We thank the entire CoCo study team for help and Marcus Wortmann from the IT department for providing support during the vaccination rollout. The CoCo Study is supported by unrestricted grants from Novartis, Gilead, Kinderherzen, and Pari. GMNB is supported by German Center for Infection Research, MSW received unrestricted funding from SARTORIUS lung research. This research was funded in part by grants from the state of Lower Saxony (14 - 76103-184 CORONA-12/20) and the Federal Ministry of Health (ZMVI1-2520COR804).

## Supplementary Material

### Methods

Following written informed consent, peripheral blood samples were obtained by venipuncture. Vaccinees for this analysis were enrolled into the CoCo Study in 2020 before vaccination (See Table 1) for detecting silent seroconversions against SARS-CoV-2 infection (29). Individuals with previous PCR confirmed SARS-CoV-2 infection or SARS-CoV-2 seroconversion before vaccinations were excluded from this analysis. Blood samples from individuals vaccinated with the BioNTech/Pfizer vaccine BNT162b2 were obtained mean 17.6 day (range 2 to 27 days) after the first and mean 21 days (range 6 to 36 days) after the second dose. Characteristic for convalescent COVID-19 patients with reverse transcriptase polymerase chain reaction (RT-PCR) test confirmed SARS-CoV-2 infection before the occurrence of VOC in Germany are summarized in Table 1.

**Table 1:** Characteristics of convalescent COVID-19 patients and BNT162b2 vaccinees.

| Reconvalescent COVID-19 patients |  |  |  |  |  |  |  |
| --- | --- | --- | --- | --- | --- | --- | --- |
|  |  |  | COVID-19 Severity |  |  |  |  |
| N = | Age<br>in years (range) | Male/female | Weeks since<br>diagnosis<br>(range) | outpatient | hospitalized | ICU |  |
| 26 | 37 (19-57) | 10/16 | 8.5 (3-36) | 25 | 1 | 0 | Fig. 3A+B |
| 20 | 54 (20-84) | 12/8 | 10.1 (7-14) | 4 | 9 | 7 | Fig. 3D |
| BNT162b2 vaccinees |  |  |  |  |  |  |  |
| 148 | 40.6 (22-66) | 63/85 | na | na | na | na |  |

After blood collection, we obtained plasma from EDTA or lithium heparin blood (S-Monovette, Sarstedt) and stored it at minus 80°C until use. PBMCs were isolated from whole blood samples by Ficoll gradient centrifugation and stored in liquid nitrogen until use. For virus neutralization assays, we incubated plasma samples at 56°C for 30 min to inactivate putative infectious virus.

### Pseudotyping of VSV and transduction experiments

Vesicular stomatitis virus (VSV)-based pseudotype particles were prepared according to a published protocol (30) making use of a replication-deficient VSV vector that does not contain the VSV-G open reading frame but instead codes for two reporter proteins, enhanced green fluorescent protein and firefly luciferase (FLuc), VSV* ΔG-FLuc (kindly provided by Gert Zimmer, Institute of Virology and Immunology, Mittelhäusern, Switzerland(31)). For pseudotype production, 293T cells expressing the desired viral glycoproteins upon transfection were inoculated with VSV* ΔG-FLuc (multiplicity of infection = 3) and incubated for 1 h at 37 °C. Thereafter, the inoculum was removed and cells were washed, before culture medium containing anti-VSV-G antibody (culture supernatant from I1-hybridoma cells; ATCC no. CRL-2700) was added and cells were further incubated. At 16-18 h post inoculation, VSV pseudotypes were harvested. For this, the culture supernatant was collected and centrifuged in order to pellet cellular debris (2,000 x g, 10 min, room temperature) and the clarified supernatants were aliquoted and stored at −80 °C until further use.

For neutralization experiments, heat-inactivated plasma samples (56 °C, 30 min) were serially diluted in culture medium. Next, equal volumes of pseudotype particles and plasma dilution (or medium without serum, control) were mixed and incubated for 30 min at 37 °C, before being inoculated onto Vero cells grown in 96-well plates. At 16-18 h postinoculation, transduction efficiency was analyzed. For this, the culture supernatant was removed and cells were lysed by incubation for 30 min at room temperature with lysis buffer (0.5 % Triton-X-100 in PBS). Next, lysates were transferred into white 96-well plates and FLuc activity was measured using a commercial substrate (Beetle-Juice, PJK) and a Hidex Sense plate luminometer (Hidex).

### Serology

SARS-CoV-2 IgG serology was performed by quantitative ELISA (anti-SARS-COV-2 S1 spike protein domain/receptor binding domain IgG SARS-CoV-2-QuantiVac; Euroimmun, Lübeck, Germany) in all individuals according to the manufacturer’s instructions. Antibody levels are expressed as RU/mL assessed from a calibration curve with values above 10 RU/mL defined as positive. Anti-SARS-COV-2 S1 spike protein domain IgA; Euroimmun, Lübeck, Germany) was done according to the manufacturer’s instructions. Antibody amounts are expressed as IgG ratio (optical density divided by calibrator); values below 0.8 were defined as negative, those between 0.8 and 1.1 were classified as intermediate, and values above 1.1 were defined as positive. The cPass Neutralization Antibody Detection kit (GeneScript) was used to detect circulating neutralizing antibodies against SARS-CoV-2 that block the interaction between the receptor binding domain (RBD) of the viral spike glycoprotein with the ACE2 cell surface receptor.

### Detection of IFNγ by SARS-CoV-2 Interferon Gamma Release Assay (IGRA)

SARS-CoV-2-specific T cell response was determined by measuring IFNγ production upon SARS-CoV-2 antigen stimulation using (SARS-CoV-2 Interferon Gamma Release Assay, IGRA, Euroimmun, Lübeck, Germany). Briefly, PBMCs were seeded at a density of 10^6^ cells/well and stimulated with manufacturer’s selected parts of the SARS-CoV-2 S1 domain of the Spike Protein for a period of 20-24 h. Negative and positive controls were carried out according to the manufacturer’s instruction. Following stimulation, supernatants were isolated through centrifugation and IFN-γ measured using IFNγ ELISA. Background signals from negative controls were subtracted and final results calculated in mlU/ml using standard curves.

### Detection of antiviral T-cell frequencies by IFNγ enzyme-linked immunospot (EliSpot) assay

Detection of SARS-CoV-2-specific T lymphocytes was achieved by IFNγ EliSpot assay as previously described (20). Briefly, PBMCs were isolated from blood samples by discontinuous density gradient centrifugation, resuspended in culture medium (CM) consisting of RPMI1640 (Lonza, Vervies, Belgium) supplemented with 10 % human AB serum (C.C.pro, Oberdorla, Germany) at a concentration of 1×10^7^ cells/ml, seeded in 24-well plates and rested overnight. Rested PBMCs were co-cultured in anti-IFNγ pre-coated EliSpot plates (Lophius Bio-sciences, Regensburg, Germany) for 16-18 h at a density of 2.5×10^5^ cells/well with specific antigens of interest. For stimulation of each sample, overlapping peptide pools against SARS-CoV-2 S protein, hCoV OC43 S protein, hCoV 229E S protein (JPT, Berlin, Germany) and CMV pp65 (Miltenyi Biotec) were used at a final concentration of 1 µg of each peptide/ml peptide pool. Cells stimulated with staphylococcal enterotoxin B (1 µg/ml, SEB, Merck, Taufkirchen, Germany) served as positive control and PBMCs incubated in media alone as negative control (NC). IFNγ secretion was detected using streptavidin-alkaline phosphatase (Mabtech Stockholm, Sweden) and revealed by 5-13 bromo-4-chloro-3-indolyl phosphate/nitroblue tetrazolium (BCIP/NBT Liquid Substrate, Merck, Darmstadt, Germany). Spots were counted using AID EliSpot 8.0 on an AID iSpot spectrum reader system (both from AID, Strassberg, Germany). Means of duplicate wells were calculated and expressed as the number of spots per well (spw). PBMCs were stained with anti-CD45 APC-H7, anti-CD3 FITC, anti-CD4 PerCP and anti-CD8 APC (BioLegend and BD Biosciences) and analysed using the FACSCanto 10c system (BD Biosciences, Heidelberg, Germany) and BD FACSDiva Software version 8.0.1 for calculation of spots/10,000 CD3+ T cells.

### Statistics

Data are presented as single results where possible with median of groups depicted as lines. Alternatively, data are depicted as mean ± standard deviation (SD). Comparison between groups were performed by two-paired student’s test, ANOVA with Bonferroni’s post hoc analysis, or Fisher’s exact test, where appropriate. Statistical analysis was performed by GraphPad Prism 5.01, which was also used for data illustration. A p-value of <0.05 was considered as significant.

## References

1. Baden LR, et al. Efficacy and Safety of the mRNA-1273 SARS-CoV-2 Vaccine. N Engl J Med. 2021;384(5):403–16.

2. Polack FP, et al. Safety and Efficacy of the BNT162b2 mRNA Covid-19 Vaccine. N Engl J Med. 2020;383(27):2603–15.

3. Voysey M, et al. Safety and efficacy of the ChAdOx1 nCoV-19 vaccine (AZD1222) against SARS-CoV-2: an interim analysis of four randomised controlled trials in Brazil, South Africa, and the UK. Lancet. 2021;397(10269):99–111.

4. Davies NG, et al. Estimated transmissibility and severity of novel SARS-CoV-2 Variant of Concern 202012/01 in England. medRxiv. 2020:2020.12.24.20248822.

5. Volz E, et al. Transmission of SARS-CoV-2 Lineage B.1.1.7 in England: Insights from linking epidemiological and genetic data. medRxiv. 2021:2020.12.30.20249034.

6. Wang P, et al. Increased Resistance of SARS-CoV-2 Variants B.1.351 and B.1.1.7 to Antibody Neutralization. bioRxiv. 2021:2021.01.25.428137.

7. Xie X, et al. Neutralization of SARS-CoV-2 spike 69/70 deletion, E484K, and N501Y variants by BNT162b2 vaccine-elicited sera. bioRxiv. 2021:2021.01.27.427998.

8. Hoffmann M, et al. SARS-CoV-2 variants B.1.351 and P.1 escape from neutralizing antibodies. Cell. 2021.

9. Wang Z, et al. mRNA vaccine-elicited antibodies to SARS-CoV-2 and circulating variants. bioRxiv. 2021:2021.01.15.426911.

10. Wu K, et al. mRNA-1273 vaccine induces neutralizing antibodies against spike mutants from global SARS-CoV-2 variants. bioRxiv. 2021:2021.01.25.427948.

11. Sahin U, et al. COVID-19 vaccine BNT162b1 elicits human antibody and TH1 T cell responses. Nature. 2020;586(7830):594–9.

12. Sahin U, et al. BNT162b2 induces SARS-CoV-2-neutralising antibodies and T cells in humans. medRxiv. 2020:2020.12.09.20245175.

13. Robertson JFR, et al. Delayed second dose of the BNT162b2 vaccine: innovation or misguided conjecture? Lancet. 2021;397(10277):879–80.

14. Gaebler C, et al. Evolution of Antibody Immunity to SARS-CoV-2. bioRxiv. 2021:2020.11.03.367391.

15. Sette A, and Crotty S. Adaptive immunity to SARS-CoV-2 and COVID-19. Cell. 2021;184(4):861–80.

16. Grifoni A, et al. Targets of T Cell Responses to SARS-CoV-2 Coronavirus in Humans with COVID-19 Disease and Unexposed Individuals. Cell. 2020;181(7):1489–501 e15.

17. Peng Y, et al. Broad and strong memory CD4(+) and CD8(+) T cells induced by SARS-CoV-2 in UK convalescent individuals following COVID-19. Nat Immunol. 2020;21(11):1336–45.

18. Murugesan K, et al. Interferon-gamma release assay for accurate detection of SARS-CoV-2 T cell response. Clin Infect Dis. 2020.

19. Petrone L, et al. A whole blood test to measure SARS-CoV-2-specific response in COVID-19 patients. Clin Microbiol Infect. 2021;27(2):286 e7–e13.

20. Bonifacius A, et al. COVID-19 immune signatures reveal stable antiviral T cell function despite declining humoral responses. Immunity. 2021;54(2):340–54 e6.

21. Becker M, et al. Exploring beyond clinical routine SARS-CoV-2 serology using MultiCoV-Ab to evaluate endemic coronavirus cross-reactivity. Nat Commun. 2021;12(1):1152.

22. Braun J, et al. SARS-CoV-2-reactive T cells in healthy donors and patients with COVID-19. Nature. 2020;587(7833):270–4.

23. Le Bert N, et al. SARS-CoV-2-specific T cell immunity in cases of COVID-19 and SARS, and uninfected controls. Nature. 2020;584(7821):457–62.

24. Mateus J, et al. Selective and cross-reactive SARS-CoV-2 T cell epitopes in unexposed humans. Science. 2020;370(6512):89–94.

25. Sette A, and Crotty S. Pre-existing immunity to SARS-CoV-2: the knowns and unknowns. Nat Rev Immunol. 2020;20(8):457–8.

26. Brockman MA, et al. Weak humoral immune reactivity among residents of long-term care facilities following one dose of the BNT162b2 mRNA COVID-19 vaccine. medRxiv. 2021.

27. Liao M, et al. Single-cell landscape of bronchoalveolar immune cells in patients with COVID-19. Nat Med. 2020;26(6):842–4.

28. Bradley T, et al. Antibody Responses after a Single Dose of SARS-CoV-2 mRNA Vaccine. N Engl J Med. 2021.

29. Behrens GMN, et al. Strategic Anti-SARS-CoV-2 Serology Testing in a Low Prevalence Setting: The COVID-19 Contact (CoCo) Study in Healthcare Professionals. Infect Dis Ther. 2020;9(4):837–49.

30. Kleine-Weber H, et al. Mutations in the Spike Protein of Middle East Respiratory Syndrome Coronavirus Transmitted in Korea Increase Resistance to Antibody-Mediated Neutralization. J Virol. 2019;93(2).

31. Berger Rentsch M, and Zimmer G. A vesicular stomatitis virus replicon-based bioassay for the rapid and sensitive determination of multi-species type I interferon. PLoS One. 2011;6(10):e25858.

